# HPV self-sampling for cervical cancer screening during wartime: a pilot implementation study from a conflict-affected region of Ukraine

**DOI:** 10.64898/2026.01.01.26343310

**Authors:** Olexiy Kovalyov, Kostyantyn Kovalyov, Olena Deinichenko, Joakim Dillner, Laila Sara Arroyo Mühr

**Author notes:** **Corresponding authors: Olexiy Kovalyov** (primary, local responsibility): Zaporizhzhia State Medical and Pharmaceutical University, Zaporizhzhia, Ukraine., **Laila Sara Arroyo Mühr** (co-corresponding, scientific responsibility), Center for Cervical Cancer Elimination, CLINTEC, Karolinska Institutet and Karolinska University Hospital, 14186, Stockholm, Sweden.

## Abstract

**Introduction:** Cervical cancer screening is difficult to initiate and sustain during war due to disrupted health services, population displacement and limited clinical capacity. Evidence on whether human papillomavirus (HPV)-based screening using self-sampling can function as a screening pathway during armed conflict is unknown. This study evaluated the feasibility, acceptability and operational performance of HPV self-sampling for cervical cancer screening in a war-affected region of Ukraine.

**Methods:** Women aged 30–60 years living in Zaporizhzhia Oblast, Ukraine, were offered HPV testing using self-collected cervicovaginal samples distributed through outpatient clinics, community outreach activities and refugee centres. Samples were analysed locally using PCR-based HPV assays with external laboratory quality assurance. Women positive for HPV16/18 were referred directly for colposcopic evaluation. Primary outcomes included screening uptake, kit return rate, HPV positivity and follow-up completion among HPV16/18-positive women. Secondary outcomes included turnaround times, HPV positivity by age and sampling setting, and participant feedback.

**Results:** Between April and December 2025, 593 women were offered HPV self-sampling; only four (0.67%) declined participation. Of 589 distributed kits, 572 samples were returned and analysed (attendance rate 96.46%, 572/596), with non-return occurring only in refugee centres. Overall, 52/572 women (9.1%) tested positive for targeted HPV types, including 22 (3.8%) positive for HPV16/18 and 30 (5.2%) positive for HPV31/33/35/45/52/58. All women testing positive for HPV16/18 were contacted and referred for specialist follow-up; among those completing follow-up, all were assessed within two months. Median turnaround time from self-sampling to results was 6.3 days. All recorded participant feedback was positive.

**Conclusion:** HPV self-sampling–based cervical cancer screening can be initiated and maintained during active war with high uptake, timely laboratory processing and effective referral of women at highest risk. Simplified, low-contact screening pathways may support cervical cancer prevention under wartime conditions.

**Trial registration:** ClinicalTrials.gov identifier: NCT07275333.

**What is already known on this topic:**

- HPV-based screening is the most effective method for cervical cancer prevention and is recommended globally.
- HPV self-sampling increases participation and acceptability in stable healthcare settings.
- Whether cervical cancer screening can be implemented during an active armed conflict is unknown.

**What this study adds:**

- Demonstrates that HPV self-sampling–based screening can be initiated and sustained during active war.
- Shows high uptake, timely laboratory processing and effective referral of HPV16/18-positive women under severe operational constraints.
- Identifies distribution models that minimise sample loss in highly mobile and displaced populations.

**How this study might affect research, practice or policy:**

- Supports inclusion of HPV self-sampling in cervical cancer prevention strategies for conflict-affected settings.
- Informs programme design choices, favouring simplified, single-visit screening pathways when population mobility is high.
- Provides implementation evidence relevant for countries rebuilding screening programmes during or after military conflicts.

## Introduction

Cervical cancer remains a major cause of morbidity and mortality despite being largely preventable.(1) Persistent infection with high-risk human papillomavirus (HPV) is a necessary cause of almost all cervical cancers, and HPV-based screening is now the preferred method globally due to its higher sensitivity and suitability for automation.(2, 3) The WHO elimination strategy has accelerated adoption of HPV testing in countries with stable healthcare systems.(4, 5)

Ukraine, however, did not have an organised cervical cancer screening programme before the war. Small pre-war projects explored HPV testing, but without population-based implementation, standardised pathways, or validated assays.(6) The escalation of armed conflict further reduced the already limited opportunities to establish screening, as clinic capacity, supply chains and population mobility deteriorated. In such conditions, traditional clinic-based screening is difficult to scale or sustain.

HPV self-sampling offers not only a more acceptable option for women but also a pragmatic operational strategy to initiate screening where access to clinics is disrupted.(7, 8) It can reach displaced populations, reduce dependency on clinical staff, and maintain screening during insecurity. However, there is very little evidence on how to integrate self-sampling into a functional screening pathway during an active war, where recruitment, transport of samples, use of locally available assays (often non-validated), quality assurance, digital reporting, and follow-up all face additional constraints.

To address this gap, we implemented an HPV-based cervical screening pathway using self-sampling in Zaporizhzhia, Ukraine, a region close to the frontline. The initiative aimed to establish a workable screening pathway under wartime conditions by recruiting women through multiple channels, using a locally available HPV assay whose analytical performance was assessed via international proficiency testing, and integrating digital communication and follow-up. This pilot study evaluates the feasibility and acceptability of this model and provides implementation lessons for cervical cancer prevention in conflict-affected settings.

## Methods

### Study design

This pilot implementation study evaluated the feasibility, uptake, HPV positivity and follow-up pathways of an HPV-based cervical screening model using self-collected samples in Zaporizhzhia, Ukraine. The study was designed as a pragmatic implementation pilot, allowing flexible operational procedures in a rapidly changing security environment. The study size was determined pragmatically based on the number of women reachable during the study period under wartime conditions, and no formal sample size calculation was performed.

Quantitative outcomes included participation, kit return rate, HPV positivity and completion of clinical follow-up. Operational challenges, workflow adaptations and contextual disruptions were prospectively documented by the implementation team as part of routine programme monitoring.

Acceptability was assessed using a structured feedback field in the clinical database. Healthcare providers recorded brief spontaneous comments from participants regarding ease of use, comfort and willingness to repeat self-sampling. These entries formed part of routine documentation and were not collected through formal qualitative interviews.

### Setting

The study was conducted in Zaporizhzhia Oblast, southeastern Ukraine, a region affected by ongoing military activity and intermittent infrastructure disruptions, where organised cervical cancer screening had not been available.

Programme preparation began in September 2024, including ethical approvals, digital platform development, documentation and staff training. Participant enrolment and HPV self-sampling started on 1 April 2025 and is still ongoing; the present analysis includes data collected up to 5 December 2025.

The programme was implemented by Zaporizhzhia State Medical and Pharmaceutical University (ZSMPhU), affiliated outpatient clinics, the World Against Cancer Foundation (Ukraine) and the International HPV Reference Center, Karolinska Institutet (Sweden). Self-sampling was selected to reduce barriers related to mobility restrictions, safety concerns and limited clinic availability.

### Participants

Women were recruited through outpatient clinics, community outreach activities and refugee centres. A small number of women in military service or volunteering roles participated when attending routine clinical care.

To minimise selection and information bias, eligibility criteria were intentionally broad to maximise access under conflict conditions. Women aged 30–60 years who were present in the region and could provide informed consent were eligible. Exclusion criteria included pregnancy, prior hysterectomy and inability to self-sample for medical or cognitive reasons. No exclusions were made based on ethnicity, displacement status or socio-economic background.

The programme targeted women eligible for cervical screening. Sex was recorded as female in routine records; gender identity variables were not collected and were therefore not analysed. Clinicians, family physicians and trained community volunteers provided information about the programme and offered women the option to self-sample either on-site or in a private setting of their choice. Participation was voluntary, and written informed consent was obtained.

### Intervention

The intervention consisted of offering HPV testing on self-collected cervicovaginal samples. FLOQSwabs® (COPAN). Self-sampling kits were distributed through outpatient clinics, refugee centres and community outreach activities, with verbal and illustrated instructions provided by trained staff.

At enrolment, healthcare staff registered participants in a secure database and completed a brief screening-history checklist to verify whether the woman had been screened within the past three years or had unresolved HPV positivity. Women with documented recent (<3 years) HPV-negative tests were not screened again.

At clinics, women who agreed to participate could choose between private self-sampling in a dedicated on-site space or taking the self-sampling kit home for later collection. In community outreach settings, volunteers provided verbal information, demonstrated the procedure and collected completed kits during the same visit. In refugee centres, kits were distributed together with instructions for private self-sampling and return through volunteers or family physicians.

After self-sampling, kits were returned to clinics or designated collection points. Clinicians and volunteers documented spontaneous participant feedback in a structured free-text field. These comments were later categorised as positive, neutral or negative using a simple AI-assisted rule-based classifier, followed by manual verification.

Women testing positive for HPV16 or HPV18 were contacted by telephone and referred for colposcopic evaluation. Women positive for the HPV types 31, 33, 45, 52 and 58 were scheduled for repeat HPV testing after 12 months to assess viral persistence.

### Laboratory procedures

All self-collected samples were transported to the DiaServis laboratory (https://diaservis.ua) in Zaporizhzhia at room temperature. A second laboratory (Diagen, https://diagen.com.ua) served as contingency capacity but was not used during the study period. On arrival, samples were registered and stored at 2–8 °C until processing.

DNA extraction was performed using the MagPurix® 12S automated system (Zinexts, Taiwan). HPV detection was conducted using two PCR-based assays routinely available in the laboratory: Sacace HPV Genotypes 14 Real-TM Quant (Italy) and Macro-Microtest 14 Types of HPV Nucleic Acid Typing Detection Kit (Fluorescence PCR; China). Both assays detect a panel of 14 HPV genotypes, including several high-risk types; however, neither assay is formally clinically validated for primary cervical screening according to WHO or international validation criteria.(9, 10)

To ensure analytic reliability, the International HPV Reference Center (Karolinska Institutet) conducted external verification.(11) A blinded proficiency panel containing known HPV-positive and HPV-negative samples was provided to DiaServis (and Diagen), and the laboratory’s results were compared with reference standards. Verification included assessment of genotype-specific detection accuracy and evaluation for potential cross-contamination (false positivity). The laboratory demonstrated proficient analytical performance for high-risk HPV detection. Proficiency testing is performed annually to ensure sustained performance over time.

Laboratory results were then transferred to Karolinska Institutet and uploaded into the secure screening database, where clinicians could view results and arrange follow-up. HPV results were reported using WHO Target Product Profile (TPP) categories, distinguishing HPV16, HPV18, and the other HPV genotypes 31, 33, 35, 45, 52, 58.(12) Genotypes outside these categories were not considered to require clinical management in a war setting.

### Outcome measures

The primary outcomes reflected the essential components of a complete screening pathway and included screening uptake, return rate of self-sampling kits, HPV prevalence, and completion of clinical follow-up among women who tested positive for HPV16/18. Screening uptake was defined as the proportion of eligible women who accepted self-sampling when offered, while the return rate reflected the proportion of distributed kits that were returned for processing. HPV prevalence was reported overall and according to WHO Target Product Profile (TPP) categories, distinguishing HPV16, HPV18 and six other oncogenic HPV types.(12) Completion of follow-up was defined as attendance at colposcopy or specialist assessment among women with HPV16/18.

Secondary outcomes included differences across recruitment channels (clinics, community outreach activities, refugee centres and military settings) in screening uptake, kit return and HPV prevalence. Additional secondary measures included the location where self-sampling occurred (clinic, home or outreach setting) and the acceptability of self-sampling. Acceptability was assessed through brief spontaneous participant feedback recorded by clinicians during routine care and subsequently categorised as positive, neutral or negative.

The study also aimed to examine feasibility under wartime conditions. Feasibility was assessed by documenting operational challenges such as disruptions to electricity or internet connectivity, delays in sample transport, and the need for adaptations (e.g., temporary manual registration when digital systems were unavailable following shelling). These contextual data allowed examination of whether self-sampling and HPV testing could be maintained reliably despite security constraints. Qualitative interviews with healthcare providers and participants are being collected and are therefore not included in this analysis.

### Statistical analysis

All analyses were primarily descriptive. Participant characteristics, recruitment pathways, screening uptake, kit return rates, HPV positivity and follow-up outcomes were summarised using counts and percentages. The study was designed to assess feasibility and implementation rather than to detect statistically significant differences or build multivariable models.

HPV positivity was reported overall and stratified by age group and place of sample collection. Age was analysed in 10-year categories corresponding to screening-relevant age groups, and HPV results were grouped according to WHO Target Product Profile categories to reflect clinically meaningful risk stratification. Exploratory comparisons of high-risk HPV positivity across age groups were performed using a chi-square test, with invalid results excluded from analysis.

Follow-up completion was calculated as the proportion of women with HPV16/18 who attended specialist assessment within the predefined two-month follow-up window.

Free-text feedback from participants was analysed using a simple AI-assisted classifier that categorised responses as positive, neutral or negative. All AI-generated classifications were manually reviewed for accuracy. Quantitative data were compiled and summarised using standard descriptive methods, and routine data validation checks ensured internal consistency.

### Ethics approval

The study was approved by the Ethics Committee of the Educational and Scientific Medical Center “Universitetskaya Klinika,” Zaporizhzhia State Medical and Pharmaceutical University, Ministry of Health of Ukraine (Protocol No. 9, 24 September 2024), and complied with the principles of the Declaration of Helsinki. Approval for quality assurance and data-related components was obtained from the Swedish Ethical Review Authority (2024-06579-01). All participants provided written informed consent prior to enrolment.

### Patient and public involvement

Patients were not involved in the design of the study, the formulation of the research questions or the selection of outcome measures. The programme was implemented under wartime conditions, which limited opportunities for formal patient and public involvement. However, the local non-governmental organisation World Against Cancer supported community awareness activities and facilitated communication with participating women during implementation. Informal participant feedback on the self-sampling procedure was routinely collected during programme delivery, although it did not lead to protocol modifications during the study period. Insights from community engagement and participant feedback will inform future adaptations and scale-up of the programme.

## Results

### Participant enrolment and sample return

Between 1 April and 5 December 2025, 593 eligible women were approached and offered HPV self-sampling through outpatient clinics (n = 503) and refugee centres (n = 86) in Zaporizhzhia Oblast. Only 4/593 (0.67%) women declined participation, and 589 self-sampling kits were distributed. Participant enrolment, self-sampling kit distribution and sample return are summarised in Figure 1.

**Figure 1:**
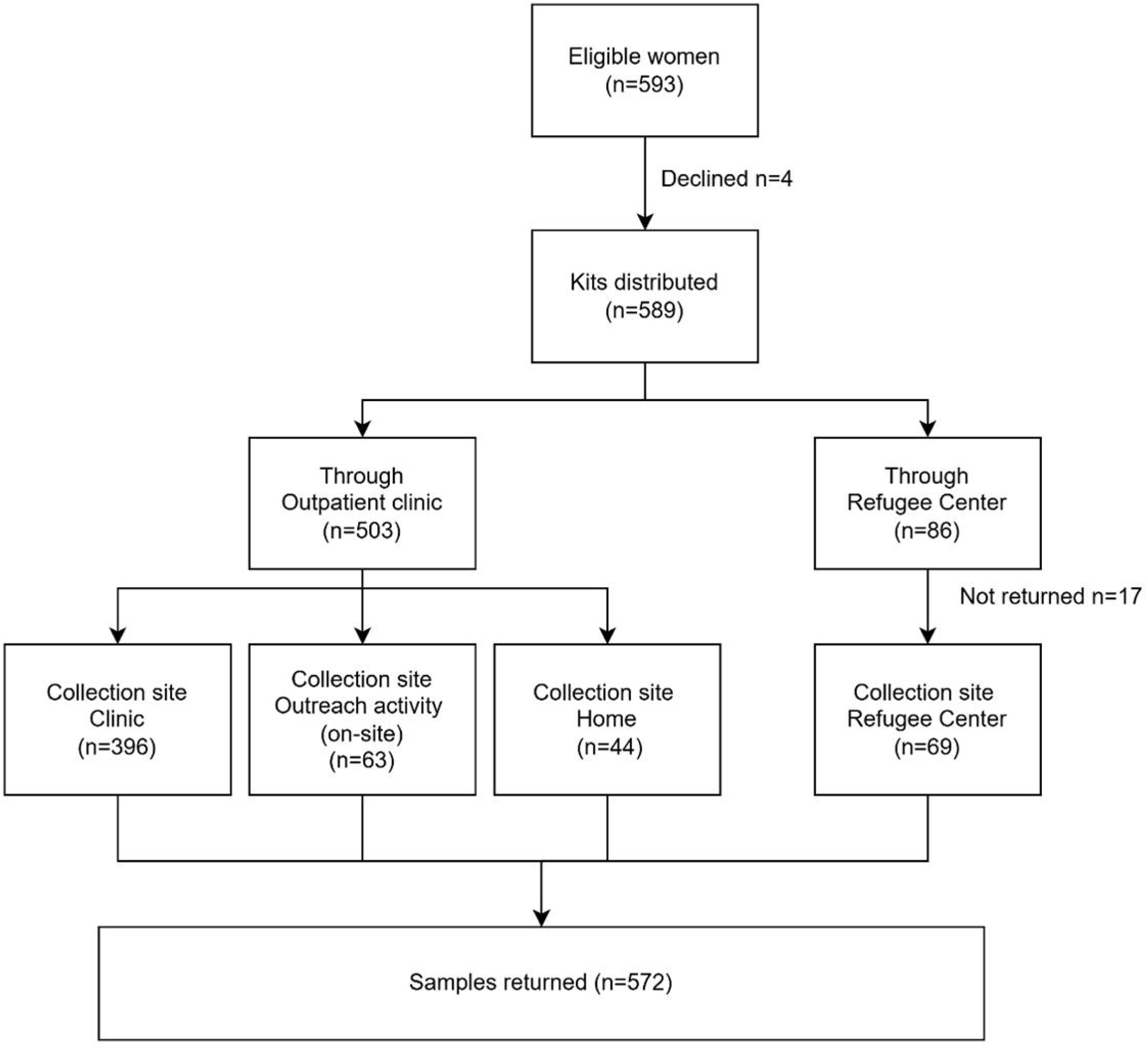
Flow of participants through the HPV self-sampling screening pathway. Flow diagram showing participant enrolment, self-sampling kit distribution through outpatient clinics and refugee centres and sample collection by setting.

Kits distributed through outpatient clinics (n = 503) included women who self-sampled on site at the clinic (n = 396), during scheduled community outreach visits (n = 63), or at home after taking the kit from the clinic (n = 44); all three modalities resulted in complete sample return. Distribution of HPV self-sampling kits at refugee centres resulted in 69 kits being returned for HPV analysis, while 17 kits were not returned (19.8% of kits distributed at the refugee centers). All kits taken home after clinic-based distribution were returned, indicating that non-return was specific to refugee-centre distribution rather than home-based self-sampling itself.

### Baseline screening history and risk stratification

Participating women were asked to complete a brief questionnaire to assess prior cervical screening history and inform baseline risk stratification. Women reporting a documented HPV-negative test within the previous three years were excluded from screening.

Among women with returned samples (n = 572), the majority (540/572) reported no prior HPV testing, while 21/572 were unsure whether they had ever been tested; these women were classified as middle risk at enrolment.

Only 11/572 women reported previous HPV testing. Among these, five reported a prior HPV-negative result obtained more than three years earlier, while four did not recall the test result, also reporting testing more than three years previously. One woman reported an HPV-positive result in 2011 but did not recall any subsequent follow-up. Another woman reported an HPV-positive result obtained two years prior to enrolment, with no follow-up documented.

Based on questionnaire responses, all women were classified as middle risk (not HPV tested) at enrolment, except for the latter participant, who was classified as a high risk due to recent HPV positivity without documented follow-up.

### HPV test results

#### HPV test results overall and by age

Among the 572 women with available HPV test results, 52 women (9.1%) tested positive for anyone of the eight targeted HPV types, including 20 positive for HPV16, 2 positive for HPV18, and 30 positive for the other HPV types 31/33/35/45/52/58. The remaining 518 women (90.6%) tested negative for these HPV types (Table 1). Two samples were classified as invalid.

**Table 1:** HPV genotype distribution by age group.

| Age | HPV Results overall |  |  |  |  |  |
| --- | --- | --- | --- | --- | --- | --- |
|  | HPV16 | HPV18 | Other HPV | Negative | Invalid | Total |
| 30-39 | 13 | 1 | 14 | 165 | 0 | 193 |
| 40-49 | 4 | 1 | 7 | 214 | 0 | 226 |
| 50-60 | 3 | 0 | 9 | 139 | 2 | 153 |
| TOTAL | 20 | 2 | 30 | 518 | 2 | 572 |

Other HPV positivity includes only HPV 31/33/35/45/52/58. Invalid results were excluded from statistical analyses.

HPV positivity varied by age group. The highest proportion of HPV-positive results was observed among women aged 30–39 years, with 28 of 193 women (14.51%) testing positive for the eight HPV types. In comparison, 12 of 226 women (5.31%) aged 40–49 years and 12 of 153 women (7.84%) aged 50–60 years tested HPV-positive. Across all age groups, HPV16 was the most frequently detected genotype (Table 1).

Excluding invalid results (n=2), HPV positivity differed across age groups (χ²=10.97, df=2, p=0.004). Positivity was highest in women aged 30–39 years (28/193, 14.5%), compared with 40–49 years (12/226, 5.3%) and 50–60 years (12/151, 7.9%).

#### HPV test results by place of sample collection

When stratified by place of sample collection, most HPV-positive results were identified among women who self-sampled at outpatient clinics, reflecting the larger number of women screened in this setting (Table 2). Among clinic-based samples (n = 459), 21 women (4.58%) tested positive for HPV16 or HPV18 and 23 (5.01%) tested positive for other high-risk HPV types.

**Table 2:** HPV test results stratified by age group and place of sample collection.

| Collection site | HPV result | Age group |  |  | TOTAL |
| --- | --- | --- | --- | --- | --- |
|  |  | 30-39 | 40-49 | 50-60 |  |
| At the Clinic | HPV16 | 12 | 4 | 3 | 19 |
|  | HPV18 | 1 | 1 | 0 | 2 |
|  | Other HPV | 13 | 5 | 5 | 23 |
|  | Negative | 132 | 167 | 114 | 413 |
|  | Invalid | 0 | 0 | 2 | 2 |
|  | Total | 158 | 177 | 124 | 459 |
| At Home | HPV16 | 1 | 0 | 0 | 1 |
|  | HPV18 | 0 | 0 | 0 | 0 |
|  | Other HPV | 0 | 1 | 3 | 4 |
|  | Negative | 11 | 18 | 10 | 39 |
|  | Total | 12 | 19 | 13 | 44 |
| At Refugee center | HPV16 | 0 | 0 | 0 | 0 |
|  | HPV18 | 0 | 0 | 0 | 0 |
|  | Other HPV | 1 | 1 | 1 | 3 |
|  | Negative | 22 | 29 | 15 | 66 |
|  | Total | 23 | 30 | 16 | 69 |
| TOTAL |  | 193 | 226 | 153 | 572 |

Other HPV positivity includes HPV 31/33/35/45/52/58. Invalid results were excluded from statistical analyses.

Among women who self-sampled at home after receiving the kit at the clinic (n = 44), five women tested positive for HPV (11.36%), most of whom (4/5) were positive for non-16/18 types.

In refugee centres, 69/86 samples were returned and analysed, with three women (4.35%) testing positive for HPV, all due to non-16/18 genotypes (Table 2).

Overall, HPV positivity was observed across all collection settings; however, numbers were small for home-based and refugee-centre sampling, and no formal statistical comparisons by setting were performed.

### Follow-up among HPV16/18-positive women

Women testing positive for HPV16 or HPV18 (n = 22) were referred directly for colposcopic evaluation according to the predefined screening pathway. All 22 women were contacted and referred for specialist gynaecologist follow-up.

By the time of data cut (5 December 2025), 11 of 22 women (50%) had completed colposcopic assessment with histological evaluation when indicated. The time from HPV test to colposcopic assessment ranged from 14 to 43 days (median 30 days, IQR 27–41), and all assessments occurred within the predefined two-month follow-up window.

Most completed follow-up examinations (8/11, 72.73%) showed no evidence of intraepithelial lesions or malignancy (NILM). Low-grade abnormalities were identified in a small number of women, including LSIL (n = 2) and ASC-US (n = 1). No high-grade lesions were identified within the available follow-up data.

Among the 11 women with pending follow-up at the time of data cut-off, 6/11 (54.55%) were still within the predefined two-month follow-up window. The remaining 5/11 (45.45%) had exceeded the planned follow-up interval, reflecting challenges related to patient mobility, security constraints and appointment attendance rather than failures in referral or clinical availability. All women with pending follow-up had been screened through outpatient clinics and have been actively recalled for colposcopic assessment.

### Operational feasibility and turnaround times

HPV self-sampling, sample transport and laboratory testing were maintained throughout the study period despite intermittent disruptions related to security conditions, electricity outages and internet connectivity. Operational adaptations, including temporary manual registration of samples and deferred digital upload of results, were implemented when required and did not result in loss of samples or test results.

Turnaround time from self-sampling to availability of HPV results in the digital platform was median 6.31 days (interquartile range [IQR] 3.10–8.32 days; range 0.24–20.26 days). The time from dispatch of samples to the laboratory to availability of results in the platform was similar, with a median of 6.03 days (IQR 3.09–8.14 days; range 0.24–19.26 days).

All women with HPV16/18-positive results who completed follow-up were assessed within the predefined two-month follow-up window. Overall, the screening pathway remained operational during the study period, demonstrating the feasibility of HPV self-sampling–based cervical screening with timely laboratory processing and follow-up under wartime conditions.

### Participant feedback and acceptability

Collection of participant feedback was not compulsory at the start of the programme; therefore, feedback was not recorded for the first 29 enrolled women. Following this initial phase, feedback entry was made a mandatory field in the digital platform and was systematically collected thereafter.

Among women for whom feedback was collected, all provided positive comments, frequently describing the procedure as good, comfortable, fast, easy, or ideal, often expressing gratitude. No negative feedback was reported.

## Discussion

The present pilot implementation study demonstrates that HPV-based cervical cancer screening using self-sampling can be initiated and maintained in a conflict-affected setting with high uptake, timely laboratory processing and appropriate referral of women at highest risk. Despite the absence of an organised screening programme prior to the war and substantial operational constraints, including population displacement, infrastructure disruptions and limited clinical capacity, fewer than 0.7% of eligible women declined participation. These findings indicate that self-sampling is both acceptable and feasible in settings where traditional clinic-based screening is difficult to implement.

At any given time, a substantial proportion of the global population lives in settings affected by war or active military conflict.(13, 14) Achieving global cervical cancer elimination therefore requires screening strategies that remain functional under wartime conditions, rather than assuming stable health systems. In this context, the present study provides rare implementation evidence showing that HPV self-sampling–based screening can be sustained during an active war, with high uptake, timely laboratory processing and effective referral of women at highest risk.

Participant feedback further supports the acceptability of self-sampling. All recorded feedback was positive, with women frequently describing the procedure as comfortable, fast and easy. While feedback was collected as part of routine care and may be subject to social desirability bias, the consistency of responses aligns with extensive evidence from stable settings demonstrating a strong preference for self-sampling.(7)The absence of negative feedback should be interpreted cautiously but nonetheless suggests high user acceptability in this context.

Screening uptake was high across recruitment channels, and HPV positivity was detected in all sample collection settings, demonstrating the ability of decentralised approaches to reach women who may otherwise remain unscreened. The overall prevalence of HPV (9.1%) and the higher positivity observed among women aged 30–39 years are consistent with patterns reported in other populations and reinforce the importance of targeting younger adult women for screening initiation.(15)

An important operational finding was the difference in kit return rates by setting and distribution model. When self-sampling was performed on site, either at outpatient clinics or during community outreach activities, or when kits were taken home after clinic-based distribution, sample return was complete. In contrast, nearly one fifth of kits distributed in refugee centres were not returned. This finding highlights the vulnerability of screening approaches that rely on delayed or unsupervised kit return in highly mobile populations, rather than a limitation of home-based self-sampling itself. The results support simplified, single-visit screening models in humanitarian and displacement settings, where population mobility and competing priorities may undermine multi-step pathways. Similar challenges have been reported in other low-resource and fragile contexts, where attrition increases when screening requires repeated contacts or delayed sample return.(16–18)

For security reasons and prioritisation of limited clinical resources, the screening pathway was intentionally designed to minimise multiple visits and triage steps. By directly referring only women positive for HPV16/18, immediate follow-up was required for just 3.8% of screened women, while women positive for HPV31/33/35/45/52/58 could safely be scheduled for later repeat testing. All women who tested positive for HPV16 or HPV18 were contacted and referred for specialist follow-up. Among those who completed colposcopic assessment within the study period, all were seen within the predefined two-month window, and no high-grade lesions were identified. Follow-up was incomplete for women diagnosed later in the enrolment period, largely reflecting population mobility and the timing of the data cut-off rather than failures in referral or care pathways. These findings demonstrate that linkage to care for women at highest risk can be achieved under wartime conditions when referral pathways are clearly defined and actively managed. At the same time, they highlight the potential value of further shortening diagnostic and treatment pathways, through rapid testing, same-day triage, or co-location of services, to minimise loss to follow-up in highly mobile and insecure settings.

Operationally, the screening pathway remained functional despite intermittent electricity and internet outages. Median turnaround time from self-sampling to availability of results in the digital platform was approximately six days, indicating that laboratory processing and reporting can be achieved within clinically meaningful timeframes even in insecure environments. Temporary adaptations, including manual registration during infrastructure disruptions, did not result in loss of samples or data, underscoring the importance of flexible, low-tech contingencies in fragile health systems.

This study has several strengths. It provides rare real-world data on cervical cancer screening implementation during active conflict, integrates external laboratory quality assurance, and evaluates the full screening pathway from recruitment to follow-up. However, limitations should be acknowledged. The study was not designed to formally compare recruitment channels or screening modalities, and sample sizes in some subgroups were small. HPV assays used locally were not formally validated for primary screening, although external proficiency testing demonstrated acceptable analytical performance. Feedback data were brief and qualitative, and more in-depth qualitative assessments are currently under preparation.

## Conclusion

This pilot demonstrates that HPV self-sampling can serve as a pragmatic entry point for cervical cancer screening in conflict-affected settings where organised programmes are absent and access to care is disrupted. Simplified, single-visit screening models, prioritisation of clinically relevant high-risk HPV types, direct referral of HPV16/18-positive women to colposcopy, and flexible operational strategies appear critical for successful implementation. As Ukraine prepares to scale cervical cancer prevention alongside the introduction of state-funded HPV vaccination, implementation approaches such as this may contribute meaningfully to progress toward cervical cancer elimination in fragile and humanitarian contexts.

## Data availability statement

Data are available upon reasonable request to the corresponding author, subject to ethical and data protection considerations.

## Ethics statements

### Ethics approval

Ethical approval was granted by the Ethics Committee of the Educational and Scientific Medical Center “Universitetskaya Klinika,” Zaporizhzhia State Medical and Pharmaceutical University, Ministry of Health of Ukraine (Protocol No. 9, 24 September 2024). Approval for quality assurance and data-related components was obtained from the Swedish Ethical Review Authority (2024-06579-01).

### Patient consent

Informed consent was obtained from all participants prior to enrolment.

### Trial registration

ClinicalTrials.gov identifier: NCT07275333.

### Author contributions

LSAM and OK conceptualised the study and secured funding. LSAM, JD, OK, KD and OD designed the study and screening pathway. OK, KK and OD coordinated field implementation, participant recruitment and data collection in Ukraine. OD oversaw clinical coordination and follow-up. Laboratory quality assurance and external verification were coordinated by LSAM and JD. LSAM conducted data analysis and interpretation. LSAM drafted the manuscript. All authors critically revised the manuscript for important intellectual content and approved the final version.

## Acknowledgements

The authors thank COPAN Italia for donating FLOQSwabs® used for HPV self-sampling. COPAN had no role in the study design, data collection, analysis, interpretation of results, or decision to submit the manuscript for publication. The authors thank the participating women, healthcare providers, community volunteers and laboratory staff involved in the implementation of the screening programme in Zaporizhzhia.

